# Rare variants found in clinical gene panels illuminate the genetic and allelic architecture of orofacial clefting

**DOI:** 10.1101/2022.08.15.22278795

**Authors:** Kimberly K. Diaz Perez, Sarah W. Curtis, Alba Sanchis-Juan, Xuefang Zhao, Taylor Head, Samantha Ho, Bridget Carter, Toby McHenry, Madison R. Bishop, Luz Consuelo Valencia-Ramirez, Claudia Restrepo, Jacqueline T. Hecht, Lina Moreno Uribe, George Wehby, Seth M. Weinberg, Terri H. Beaty, Jeffrey C. Murray, Eleanor Feingold, Mary L. Marazita, David J. Cutler, Michael P. Epstein, Harrison Brand, Elizabeth J. Leslie

**Affiliations:** Department of Human Genetics, Emory University School of Medicine, Atlanta, GA; Center for Genomic Medicine, Massachusetts General Hospital, Boston, MA 02114, USA; Program in Medical and Population Genetics, Broad Institute, Cambridge, MA 02142, USA; Department of Neurology and Surgery, Massachusetts General Hospital and Harvard Medical School, Boston, MA; Department of Biostatistics and Bioinformatics, Rollins School of Public Health, Emory University, Atlanta, GA; Agnes Scott College, Decatur, GA; Center for Craniofacial and Dental Genetics, Department of Oral and Craniofacial Sciences, University of Pittsburgh School of Dental Medicine, Pittsburgh, PA; Fundación Clinica Noel, Medellin, Colombia; Department of Pediatrics, McGovern Medical, School and School of Dentistry, UT Health at Houston, Houston, TX 77030, USA; Department of Orthodontics, University of Iowa, Iowa City, IA; Department of Health Management and Policy, University of Iowa, Iowa City, IA; Department of Epidemiology, Johns Hopkins Bloomberg School of Public Health, Baltimore, MD; Department of Pediatrics, University of Iowa, Iowa City, IA; Department of Human Genetics, University of Pittsburgh School of Public Health, Pittsburgh, PA

## Abstract

**Purpose:** Orofacial clefts (OFCs) are common birth defects including cleft lip (CL), cleft lip and palate (CLP), and cleft palate (CP). OFCs have heterogeneous etiologies, complicating clinical diagnostics as it is not always apparent if the cause is Mendelian, environmental, or multifactorial. Sequencing is not currently performed for isolated or sporadic OFCs, so we estimated the diagnostic yield for 418 genes in 841 cases and 294 controls.

**Methods:** We evaluated 418 genes using genome sequencing and curated variants to assess their pathogenicity using American College of Medical Genetics criteria.

**Results:** 9.04% of cases and 1.02% of controls had ‘likely pathogenic’ (LP) variants (p<0.0001), which was almost exclusively driven by heterozygous variants in autosomal genes. CP (17.6%) and CLP (9.09%) cases had the highest yield while CL cases had a 2.80% yield. Out of 39 genes with LP variants, nine genes, including *CTNND1* and *IRF6*, accounted for more than half of the yield (4.64% of cases). Most variants (61.8%) were ‘variants of uncertain significance’ (VUSs), occurring more frequently in cases (p=0.004), but no individual gene showed a significant excess of VUSs.

**Conclusion:** These results underscore the etiological heterogeneity of OFCs and suggest sequencing could reduce the diagnostic gap in OFCs.

## INTRODUCTION

Orofacial clefts (OFCs) are etiologically heterogeneous structural birth defects^1^, including cleft lip (CL), cleft palate (CP), and cleft lip and palate (CLP). Genetic factors, such as point mutations, copy number variants, and chromosomal abnormalities, contribute to the etiology of OFCs, especially in Mendelian OFC syndromes, which contain other structural anomalies, cognitive anomalies, or intellectual disabilities. Hundreds of rare Mendelian syndromes involving OFCs have been described but most OFC cases occur as apparently isolated birth defects (often termed non-syndromic). These are considered etiologically complex disorders with genetic and environmental risk factors. It is possible, however, that both syndromic and isolated cases have an etiology caused by genetics, environment, or the combined effect of both. The heterogeneity of OFCs is further compounded by phenotypic heterogeneity, incomplete penetrance, and variable expressivity, making clinical diagnostics challenging. Recurrence risk estimates vary with an approximate sibling recurrence of ∼4%^2^, which is much lower than the sibling risk for an autosomal dominant disorder or the empirical risk of being affected with an incompletely penetrant disorder, but is also significantly higher than expected if the risk were driven by *de novo* variants alone. It is therefore important to determine the cause of OFCs as it could inform recurrence risk estimates and the approaches for genetic counseling and clinical management.

Molecular diagnoses from genetic testing is an alternative to diagnosis based on the phenotype alone. However, genetic testing is not routinely performed for most individuals with OFCs, which are typically isolated and without any family history. Furthermore, existing commercial clinical testing panels are highly variable in their content, leading to potentially missed diagnoses. Multiple studies have explored the use of sequencing to improve diagnostics for OFCs. We previously investigated the proportion of isolated OFC cases attributable to variants in *IRF6*; however, our estimate of 0.2-0.4% was too low to recommend broad screening of this gene^3^. The use of exome sequencing (WES) and genome sequencing (WGS) in OFCs has recently increased. Basha et al. tested the diagnostic rate from WES in 46 multiplex OFC families, finding 10% of cases carried ‘likely pathogenic’ (LP) variants, primarily in genes causing autosomal dominant OFC syndromes^4^. However, WES in large cohorts has not been performed so diagnostic yield estimates in OFCs are still uncertain.

We aimed to estimate the diagnostic yield of 418 genes associated with OFCs using WGS in 841 OFC cases and 294 controls. We previously investigated *de novo* variants in 756 OFC trios from this same cohort and found 6% of sequenced trios had a *de novo* variant in genes broadly associated with OFCs^5^. Two genes (*IRF6* and *TFAP2A*) mutated in OFC syndromes were individually associated with OFCs, raising the question of the clinical impact of *de novo* variants and other types of variants in similar genes. We, therefore, developed the present study to utilize WGS to fully characterize the clinical impact of variants in OFC cases by analyzing *de novo* and transmitted single nucleotide and structural variants.

## SUBJECTS AND METHODS

### Study Population

The case sample consisted of 841 total OFC cases (765 case-parent trios, 60 parent-child dyads, and 16 singletons) sequenced through the Gabriella Miller Kids First (GMKF) Pediatric Research Program. The case sample was sequenced in three cohorts based on recruitment site/ancestry: 1) “Europeans” from the United States, Argentina, Turkey, Hungary, and Spain; 2) “Latinos” from Colombia; and 3) “Asians” from Taiwan (Supplemental Table 1). Participant recruitment occurred over many years using different research protocols, but each protocol generally included a physical exam to exclude individuals with major anomalies or known intellectual disability indicative of an OFC syndrome. This cohort is therefore enriched for isolated OFCs and depleted of multiple congenital anomalies and severe manifestations of syndromes. The case sample includes probands with cleft lip only (e.g., CL, cleft lip and cleft alveolus) (107 cases), cleft lip and cleft secondary palate (CLP; 660 cases), and cleft secondary palate only (CP; 74 cases). The cases were primarily male (56% CL, 65% CLP, 53% CP) reflecting the large proportion of the cases having CLP where males are overrepresented. The 756 trios were analyzed previously for *de novo* variants only^5^.

A total of 621 probands were considered “simplex” as they reported no family history, defined as not reporting any affected relative within the 3^rd^ degree. 220 probands reported having at least one affected relative (1^st^, 2^nd^, or 3^rd^ degree) were classified as “multiplex” (Supplemental Table 2); this included 63 probands with at least one affected parent. All probands were confirmed to be unrelated with kinship calculations using KING.

The control sample was comprised of 294 child-parent trios from the 1000 Genomes Project (1KGP)^6^. Because some 1KGP samples are derived from cell lines, these samples have excessive numbers of *de novo* variants acquired through multiple passages and are not comparable to the pattern of variation in the GMKF samples^7^. Therefore, we selected 1KGP trios through a quality control process (described below) to have approximately the same amount of total variation including *de novo* variant rates (all selected 1KGP trios had fewer than 138 *de novo* events per trio) as the case cohorts. This cohort included trios from multiple ancestries: 84 African, 116 US and European, 52 East Asian, and 42 South Asian trios. Although phenotype information is unavailable for 1KGP, we would expect at most 1 OFC in the 294 trios (882 total individuals) based on the prevalence rate of OFCs at 1 in 1000 individuals worldwide. Thus, 1KGP can serve as a comparison group as it is unlikely to have LP variants influencing risk to OFCs.

### Sequencing and Quality Control

Sequencing and variant calling of the OFC cohort was described in Bishop et al.^5^. Sequencing of the control cohort was described in Byrska-Bishop et al.^8^. The same quality control procedures were performed on case and control VCF files. We retained genotype calls with a genotype quality ≥20, read depth ≥10, and biallelic variants passing VSQR with a Quality Normalized by Depth (QD) score >4 using VCFTools (v0.1.13) and BCFtools (v1.9). Variants with >2 Mendelian errors, >5% missingness, or deviations from Hardy-Weinberg equilibrium (p<10^−7^) in unaffected samples were dropped. For *de novo* variants, we required an allele balance between 0.3 – 0.7 in each proband and < 0.05 in both parents.

### Selection of Gene List

We created a comprehensive set of 418 genes (Supplemental Table 3, Supplemental Figure 1A) to prioritize variants possibly associated with OFCs from four sources (all downloaded September 4^th^, 2020): 1) the National Health Service (NHS) Genomic Medicine Service cleft panel (v2.2), an expert-curated list of genes for familial cleft lip and/or cleft palate (CL/P), isolated and syndromic clefting; 2) the Prevention Genetics CL/P clinical genetic testing panel; 3) Clinical synopses/genes from the Online Mendelian Inheritance in Man (OMIM) that included OFCs with a known inheritance and molecular basis. OMIM clinical synopses search terms included: “cleft lip,” “cleft palate,” “oral cleft,” “orofacial cleft,” and “cleft lip and/or palate;” 4) a manually curated list from recent OFC genetic studies. The NHS panel included an evidence level indicator corresponding to expert consensus for genes on the panel: “green” for genes of known clinical utility and scientific validity, “amber” for moderate evidence levels, and “red” indicating little evidence^9^. We classified genes based on the mechanism by which variants lead to OFC phenotypes and hereafter refer to these genes as autosomal dominant (AD), autosomal recessive (AR), or X-linked (XL). Genes in which variants have been described as acting in dominant and recessive manners or unspecified modes of inheritance were considered in both AD and AR analyses. Average read depth for each gene was comparable between cases and controls and between all case populations (Supplemental Figure 1B). In our previous work, we analyzed 336 genes associated with OFCs, which included genes from OMIM and those nominated by linkage, candidate gene, and association studies. The current list of 418 genes includes 200 genes not analyzed previously, the majority of which came from NHS and OMIM. There were 118 genes on the Bishop et al. gene list absent from this analysis, most of which were GWAS genes that lacked the necessary support to be included in a clinical gene panel.

### SNV and Indel Annotation and Variant Filtering

Variants were annotated using ANNOVAR (version 201707) and Variant Effect Predictor (VEP, release 102, 103, 106). Protein-altering variants were extracted for the 418 genes. Variants were filtered using a maximum allele frequency (AF) threshold of 0.1% for variants in autosomal dominant and X-linked genes and 0.5% for variants in autosomal recessive genes using gnomAD (v2 and v3)^10^ and ExAC (v0.3)^11^, and a cohort allele count (AC) ≤ 10. Variant-level annotations used in the prioritization and interpretation included nine *in silico* pathogenicity predictions (e.g., SIFT^12^, PolyPhen^13^, MutationTaster^14^, LRT^15^, FATHMM^16^, PROVEAN^17^, MetaSVM^18^, MetaLR^18^, M-CAP^19^), CADD^20^ scores, variant pathogenicity classifications from ClinVar^21^, and constrained regions within genes^22^.

### Structural Variants (SVs) Identification and Filtering

We detected SVs in the OFC cohort with the GATK-SV discovery pipeline as previously described^23^. GATK-SV (https://github.com/broadinstitute/gatk-sv) relies on an ensemble approach that harmonizes SV detection from multiple tools, including Manta^24^ and Gatk-gCNV^25^, followed by machine learning to remove likely false positive events and then performs joint genotyping and refined variant resolution. The derived VCF file was annotated by svtk^26^ to predict the functional impact of SVs and compare AF against gnomAD SV (v2.1)^23^. We obtained SVs overlapping the 418 genes and filtered the SVs by AF (OFC cohort AF ≤ 0.03 and gnomAD SV AF ≤ 0.01). SVs overlapping recurrent genomic disorder regions were investigated independently and we reported the gene(s) within our gene list from those regions. Further inheritance-specific genotype and frequency filters were applied to identify *de novo* (gnomAD SV AF ≤ 0.001, AC ≤ 10 and cohort sample count ≤ 5), homozygous (homozygous AC ≤ 10 and absent in unaffected individuals in cohort), compound heterozygous and X-linked recessive SVs.

We also considered SVs inherited from unaffected parents if the cohort AC was ≤ 10, of which ≤ 5 were unaffected individuals. Candidate SVs (Supplemental Table 4) were manually reviewed and visually inspected in normalized read-depth plots using Integrative Genomics Viewer^27^.

### Classification into the Tier System

Rare SNVs and indels located within the 418 genes were classified into a ranked tier system designed to minimize the number of variants undergoing manual American College of Medical Genetics and Genomics (ACMG) review while retaining as many potential LP variants as possible. Each tier was based on gene or variant annotation criteria, including variant type and gene constraint (Supplemental Figure 2A). Tiers were ranked based on qualitative assessments of their likelihood to contain LP variants. Assessments were made by KDP, MRB, and EJL, and the final tiers were formed from a consensus of these assessments.

After sorting variants into tiers, we identified a cutoff point above which variants would be manually reviewed according to ACMG criteria. To determine the cutoff point, we extracted variants in ClinVar from 418 genes classified as either pathogenic (‘likely pathogenic’ or ‘pathogenic’) or benign (‘likely benign’ or ‘benign’). We sorted the 526 pathogenic and 274 benign variants into tiers (Supplemental Figure 2B, and identified Tier 1B as a point where 95% of pathogenic variants but only 49% of benign variants would be retained for review.

### ACMG Variant Classification

All SNVs and indels meeting the Tier 1B threshold, in-frame indels, and SVs meeting the review criteria were assessed using ACMG criteria blinded to case-control status^28^ (Supplemental Figure 2A). We considered variants with “damaging” pathogenicity predictions from ≥ 5 out of 9 algorithms to meet the PP3 criteria, while variants with ≥ 5 out of 9 “tolerant” predictions met the BP4 criteria (Supplemental Table 5). For criteria based on AF alone (PM2, BS1), we used the maximum AF observed in any population across gnomAD v2 genomes and exomes, gnomAD v3 genomes, and ExAC exomes. Variants with an AF < 0.001% met criteria PM2, and variants with AF ≥ 0.005% (heterozygous) and AF ≥ 0.2% (homozygous) met BS1. We estimated the maximum credible AF for a variant considering an OFC prevalence of 1 in 1,000, 5% of allelic heterogeneity, 100% of genetic heterogeneity, and 50% penetrance^29^. All variants classified as ‘pathogenic’ or ‘likely pathogenic’ were counted towards the diagnostic yield calculation and are referred to as ‘likely pathogenic’ (LP) throughout the manuscript.

### Statistical Analysis

Statistical tests were performed to calculate differences between groups using two-sided chi-square and Fisher exact tests, which were conducted using R (version 3.6.3). We performed 10,000 permutations for the chi-square tests comparing cases and controls (overall, by cleft subtype, population, and sex) to adjust for multiple hypothesis testing under the null hypothesis of no association between the number of individuals with LP variants and case-control status. The significance level was set at P < 0.05 for these tests. Odds ratios and 95% confidence intervals were estimated through chi-square tests in R.

We tested gene-based associations in genes with VUSs using the Optimal Sequence Kernel Association test (SKAT-O), which unites the Sequence Kernel Association test (SKAT) and the burden test to maximize statistical power while allowing for variants of opposite effects^30^. Data were converted to binary PLINK files and imported into the SKAT package (version 2.0.1)^31^ in R (version 3.6.3). First, we performed SKAT-O tests for 139 genes with more than one VUS or LP variant. We then excluded “solved” cases and controls with LP variants and conducted SKAT-O tests for 129 genes with more than one VUS in the remaining samples. We used a Bonferroni correction to adjust for multiple testing.

## RESULTS

We identified 2,549 SNVs, small indels, and SVs within the 418 genes from 841 OFC cases and 294 controls. After sorting variants into tiers designed to prioritize variants, we narrowed our list to 1,483 variants for manual review under ACMG criteria (Supplemental Figure 2C). On average, we reviewed 1.33 variants per case and 1.24 variants per control (p=0.07).

After ACMG review, 79 variants (5.33%) were classified as ‘likely pathogenic’ (LP) (Supplemental Table 6). The LP variants were dominated by those presumed to be loss-of-function (LoF): 46.8% were stop-gain, frameshifting indels, and canonical splice site variants; 15% were SVs. Overall, 9.04% of cases and 1.02% of controls had LP variants (p<0.0001, Figure 1). Stratifying our gene list by the mode of inheritance, we found LP variants were almost exclusively in autosomal dominant genes (8.80% of cases vs. 1.02% of controls; p<0.0001). Consistent with previous analysis of an excess of *de novo* variants in clinically relevant genes among OFC cases^5^, 3.69% of cases (vs. none in controls; p=0.0008) had a *de novo* LP variant. Notably, we did not identify any LP homozygous or compound heterozygous variants in autosomal recessive genes. This lack of signal was unexpected because a subset of the trios came from consanguineous families from Turkey and Colombia. Similarly, there was a limited contribution from X-linked genes. Only two individuals (0.24% of cases) had LP variants on the X chromosome: a hemizygous male with a LoF variant in *PHF8* inherited from his unaffected mother and a heterozygous female with a *de novo* in-frame deletion in *FLNA*.

**Figure 1.**
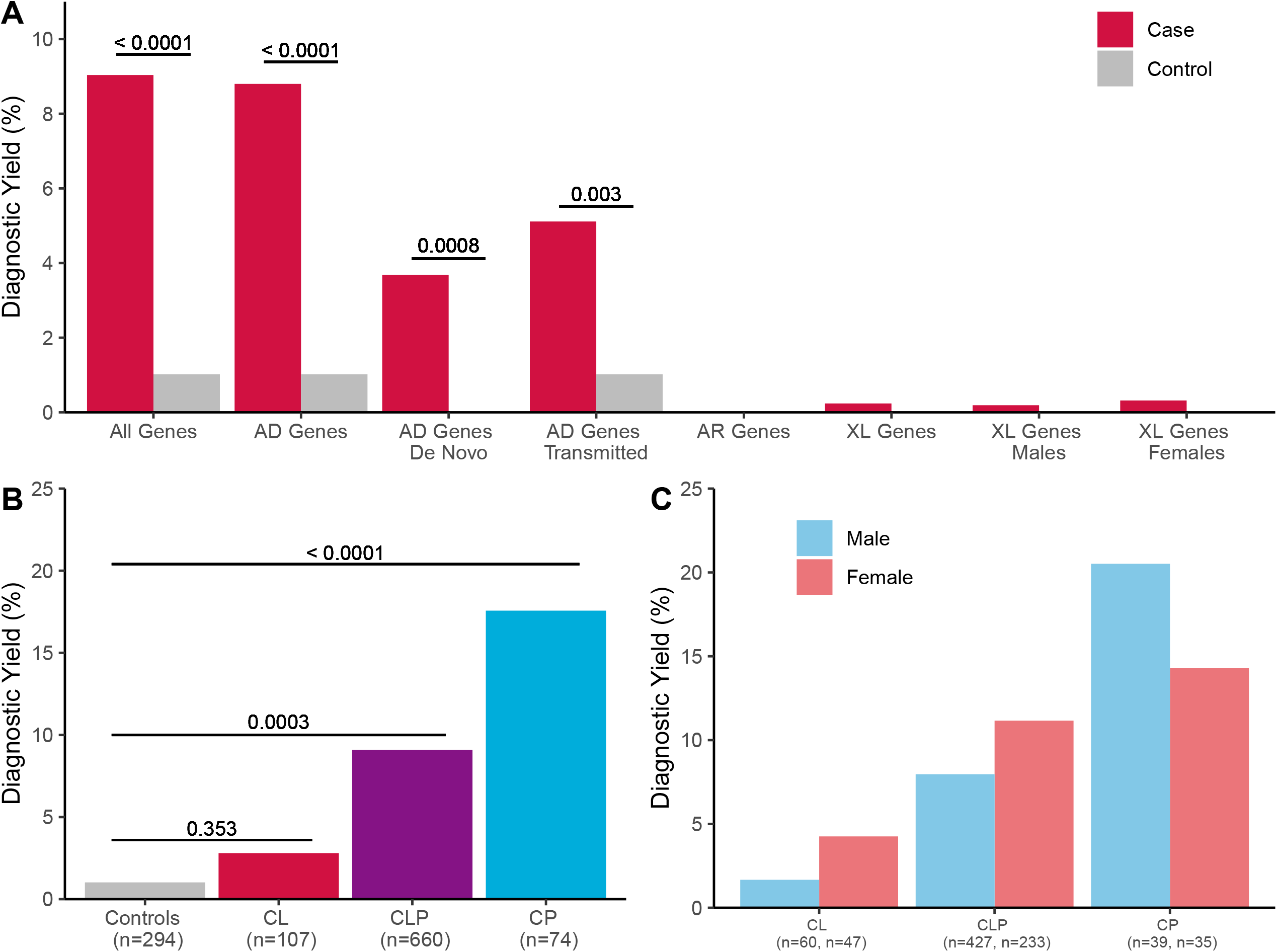
Diagnostic Yield of 418 OFC Genes. (A) The percentage of cases (red) and controls (gray) with at least one ‘likely pathogenic’ variant by mode of inheritance. (B) The percentage of individuals with ‘likely pathogenic’ variants in controls (gray) and in cases by OFC subtype: cleft lip (CL, red), cleft lip and palate (CLP, purple), and cleft palate (CP, blue). (C) Percentage of cases with a ‘likely pathogenic’ variant stratified by cleft type and proband sex (female (pink) and male (blue)). Yields were not significantly different between males and females in any cleft subtype. P-values were calculated using chi-square tests with 10,000 permutations in R.

Epidemiology and association studies suggest some differences in the genetic architecture of specific OFC subtypes^32,33^. Therefore, we stratified the case cohort to test for differences in diagnostic yield across CL, CLP, and CP subtypes (Figure 1B). Among CLP cases, which comprise 78% of the OFC cohort, 9.09% had a LP variant (60 out of 660 total CLP cases) (p=0.0003 vs. controls). The diagnostic yield was much higher among CP cases (13 out of 74 total CP cases), where 17.6% had a LP variant (p<0.0001 vs. controls and p=0.035 vs. CLP). Equally striking was the difference between CL and CLP, which have historically been viewed as a variation in severity of the same disorder and are commonly analyzed together. Only 2.80% of CL cases (3 out of 107 total CL cases) had a LP variant, which was not significantly different than controls (p=0.353) and only nominally different from CLP (p=0.045). These data suggest the differences in genetic architecture between CL and CLP seen in GWAS studies^34,35^ may extend to rare variants.

There are characteristic sex biases in OFCs where CP occurs twice as frequently in females than males, and CL/P occurs twice as frequently in males than females^36^. We considered whether these sex biases were also reflected in the yields. Although the less frequently affected sex had consistently higher diagnostic yields within each subtype, none was statistically significant (Figure 1C). These results could be consistent with a “protective effect” model; when there are disease prevalence differences between sexes, affected individuals among the less commonly affected sex have, on average, greater enrichment for disease-causing alleles or alleles of larger effect than members of the more commonly affected sex. This would also be consistent with the observation that sex biases are not as commonly observed in Mendelian OFC syndromes. We also observed small (but non-significant) differences in yield when stratifying by population by cleft type (Supplemental Figure 3). Although this is loosely correlated with OFC prevalence rates^37^, it is more likely that these differences are due to disparities in representation of these populations in reference databases that impact the filtering of variants based on AF.

The 76 LP variants in OFC cases were found across 39 genes, constituting 9.33% of the gene list (Figure 2). Sixteen genes had multiple variants, and nine of these had at least three LP variants in cases. These nine genes: *CTNND1* (6 cases), *ARHGAP29* (5 cases), *COL2A1* (5 cases), *IRF6* (5 cases), *TFAP2A* (5 cases), *CDH1* (4 cases), *CHD7* (3 cases), *PDGFC* (3 cases), and *TBX1* (3 cases, all 22q11.2 deletions) accounted for 4.64% of OFC cases alone. All of these genes with the exception of *PDGFC* (and 35 out of 39 genes with LP variants) were genes in which variants cause disease in an autosomal dominant manner. Of 163 genes associated with autosomal dominant disease, 21.5% had at least one LP variant, demonstrating the genetic heterogeneity of OFCs. Although previous studies have found LP variants in two or more disease loci in the same individual^38^, we did not identify cases with more than one LP variant.

**Figure 2.**
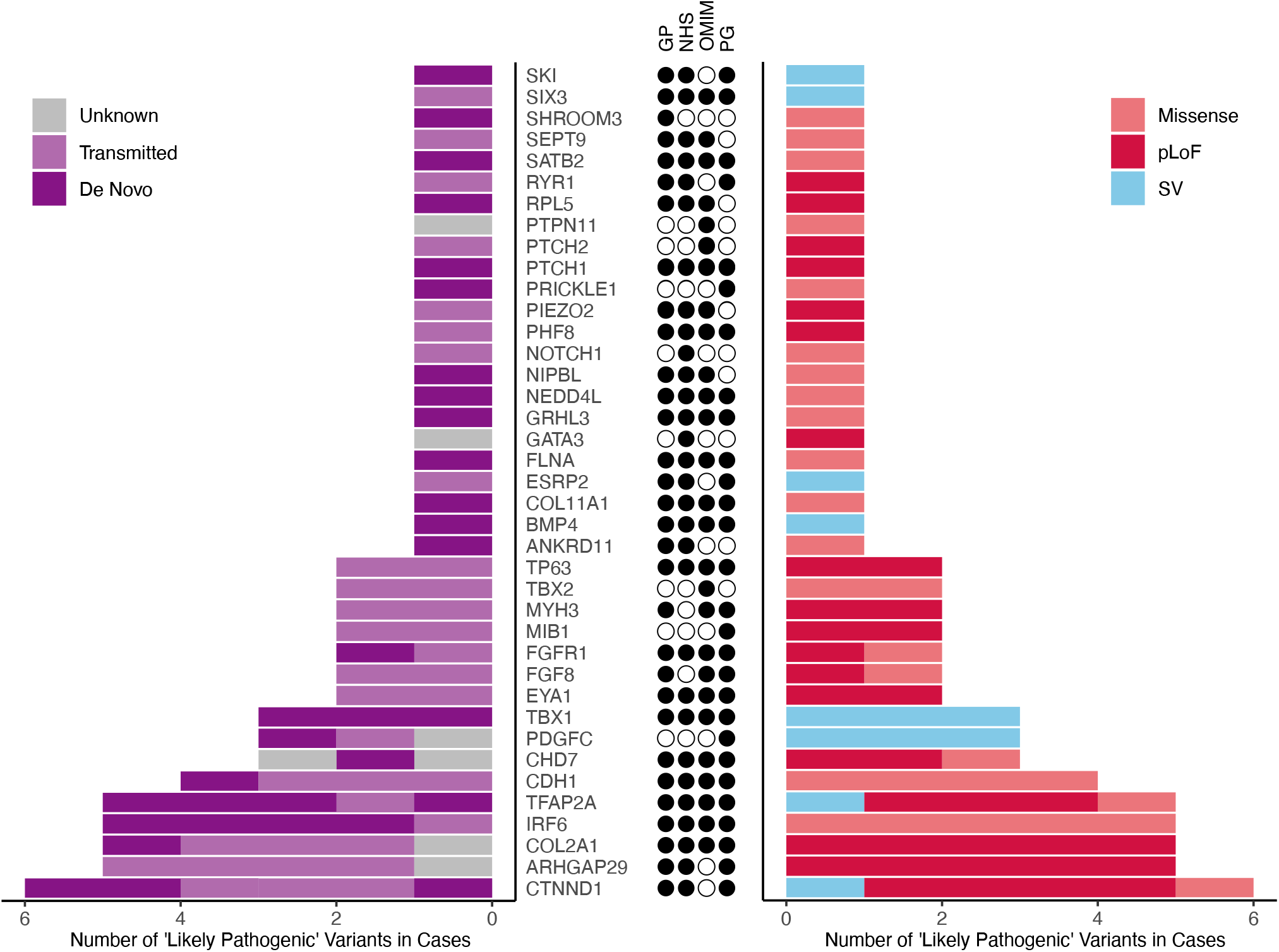
Genes with Likely Pathogenic Variants in Cases. The count of ‘likely pathogenic’ variants in cases in each of the 39 genes with ‘likely pathogenic’ variants. Individual variants are in the same order in both panels and are colored in the left panel based on the mode of inheritance: *de novo* (dark purple), transmitted (light purple), or were unknown (gray), and in the right panel based on variant consequence: missense (red) variants, predicted loss-of-function (pLoF, dark red), and structural variants (blue). The center panel shows a filled circle for each source list that each gene was on: GP (curated gene panel), NHS (NHS PanelApp list), OMIM, and PG (Prevention Genetics panel).

Previous OFC studies report incomplete penetrance for several genes, including *CTNND1 and TP63*^4,39^, but few have studied large datasets drawn from both simplex and multiplex families, allowing us to weigh the contribution of *de novo* and transmitted variants and estimate penetrance for autosomal dominant variants. A total of 220 probands were from multiplex families, defined as having at least one other affected relative (up to the 3^rd^-degree). There was no difference in yield between individuals from multiplex and simplex families (11.8% multiplex vs. 7.73% simplex, p=0.089). However, there were notable differences in the types of variants identified (Supplemental Figure 4). Twenty of the 26 LP (76.9%) variants in individuals from multiplex families were transmitted (Supplemental Figure 5); the rest were *de novo* (Supplemental Figure 6). In contrast, 52.1% (25 out of 48) of LP variants in simplex families were *de novo*, which were confirmed by visual inspection of aligned reads (Supplemental Figure 6). In three of the six families, one parent was affected, and we cannot exclude the possibility of mosaicism in other tissues. But it is also possible these variants are not the only variants conferring risk for OFCs.

Among transmitted variants in multiplex families, we asked how often the variant was transmitted by the parent with a personal or family history of OFC. We found 82.4% (14 out of 17) of variants were transmitted by the parent with a family history but found no differences in transmission for those with a maternal family history compared to a paternal family history. Of the 14 variants transmitted by a parent with a family history, 64% (9 out of 14) were transmitted by an affected parent. We can therefore estimate the penetrance for these transmitted variants in multiplex families to be 64%. If we count all transmitting parents, including unaffected parents from simplex families, the penetrance of transmitted variants falls to 25% (9/36). Interestingly, most of these variants are predicted to be loss-of-function, and impacted genes included *ARHGAP29, CTNND1*, and *TP63*, which are considered haploinsufficient with reduced penetrance^40,41^.

The majority (61.8%) of classified variants were variants of uncertain significance (VUSs). Overall, we found a significant enrichment of VUSs among OFC cases compared to controls (56.6% cases vs. 46.6% controls, p=0.004). This result was consistent across populations but not OFC subtypes (Supplemental Table 7). VUSs were not clustered in cases with LP variants or in cases without such variants as removing “solved” cases/controls resulted in a similar enrichment: 56.1% of 765 cases had at least one VUS vs. 46.7% of 291 controls (p=8.03 × 10^−3^).

One possible hypothesis to explain the excess of VUSs among cases is that there are cryptic LP variants among this set of VUSs, where some are truly disease-causing but are not recognized as such in the ACMG classification because of lack of sufficient variant- or gene-specific information. For both cases and controls, VUSs were overwhelmingly missense variants, which is not surprising given the challenges of interpreting missense variation. We expected VUS among cases would have greater “damaging” prediction scores, but there was no difference in the distribution of pathogenicity predictions aggregated under nine different algorithms (see Supplemental Table 5, Supplemental Figure 7).

We next hypothesized that the excess of VUSs in cases could be localized to genes with LP variants. Collectively, VUSs were similarly enriched among genes with LP variants (OR 1.63, p=0.008) as they were among genes without LP variants (OR 1.36, p=0.033) (Figure 3A). To further parse which sets of genes were contributing to the VUS signal, we used the evidence level of genes on the NHS panel, corresponding to three levels of support for the genes’ association with OFCs reviewed by an expert panel (“green” for high evidence, “amber” for moderate evidence, and “red” for low evidence). The enrichment of VUS was strongest among 69 “green” genes (OR 2, p=1.36 × 10^−4^) (Figure 3A). We then performed SKAT-O tests for autosomal genes with LP variants and/or VUS to pinpoint individual genes with significant VUS contributions. Although no gene reached formal significance due to an unbalanced sample size favoring cases, *PRICKLE1* (MIM: 608500) was nominally significant with an odds ratio indicating an increased risk for OFC (Supplemental Figure 8). Furthermore, many genes with multiple LP variants had an increase in odds ratio from the addition of VUSs (Figure 3B).

**Figure 3.**
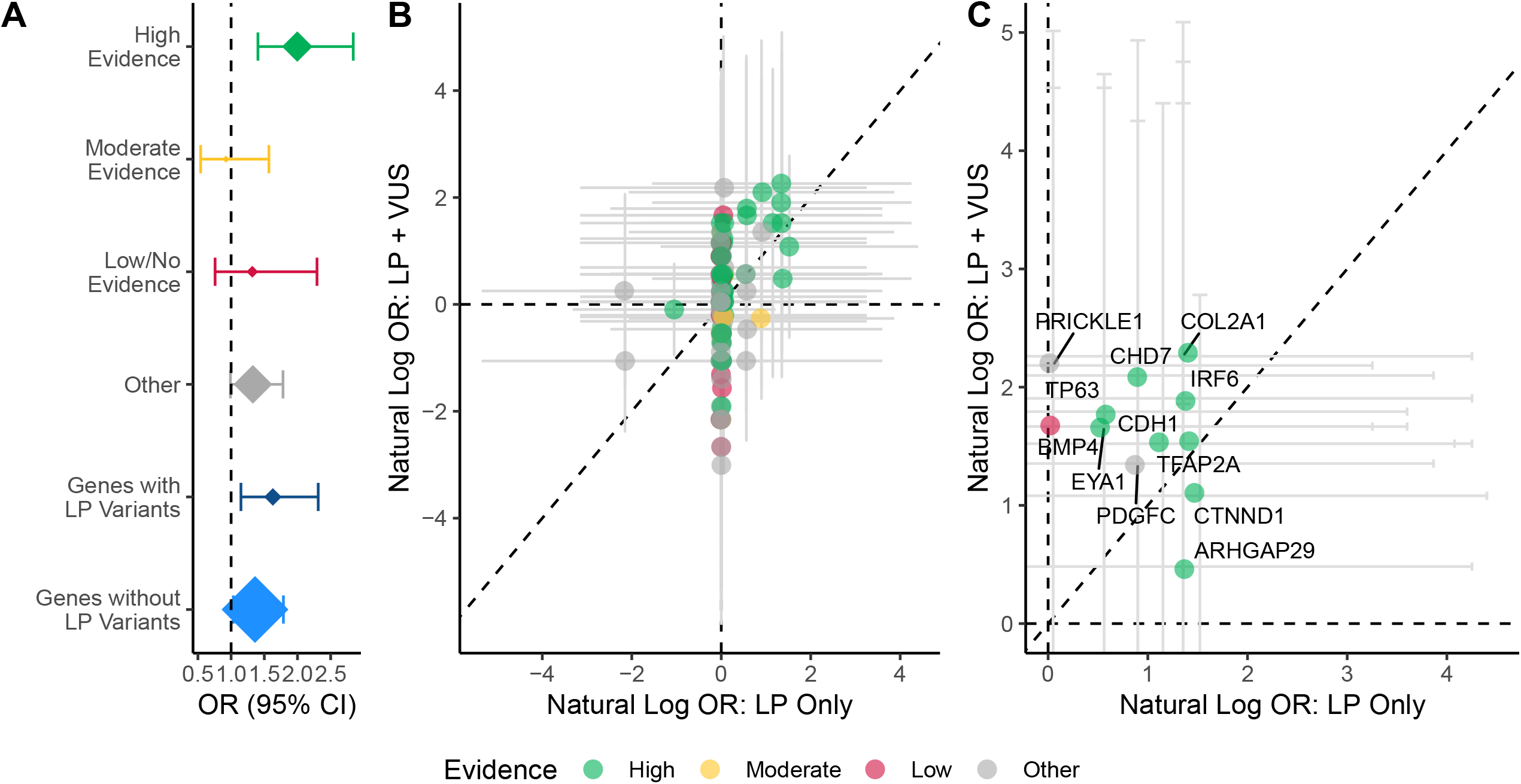
Variants of Uncertain Significance (VUSs) are enriched in cases vs. controls. (A) Enrichment of VUSs in cases for sets of genes. Odds ratios (OR) and 95% confidence intervals (95% CI) are calculated from a chi-square test. Point estimates are scaled by the number of genes in the gene set. (B) The natural log odds ratios for individual genes for ‘likely pathogenic’ (LP) variants only vs. ‘likely pathogenic’ and variants of uncertain significance (LP + VUS). (C) Individual genes with natural log odds ratios greater than 1 for ‘likely pathogenic’ (LP) variants only or likely pathogenic’ and variants of uncertain significance (LP + VUS). The dotted lines show OR = 1 for the x and y axes and y=x. In all panels, genes/gene sets are colored by the level of supporting evidence as recorded in the UK NHS Cleft Lip and Palate PanelApp: high evidence (green), moderate evidence (amber), low evidence (red), or not on the PanelApp gene list (gray).

Lastly, we assessed which gene list resulted in the highest yield of LP variants. The yield for individual lists was between 6.18% and 7.61% but no list had a statistically different yield from the others (Supplemental Table S8). The manually curated, Prevention Genetics, and NHS gene lists had the highest proportion of genes with LP variants, which is not surprising given that 82% of the 418 genes appear on at least one of those three lists (Supplemental Figure 9A). Most autosomal dominant genes with LP variants (27 out of 35) were shared among at least these three sources (Supplemental Figure 9B). In contrast, the OMIM list had the lowest percentage (10.3%) of LP variants and a yield of 6.18% for all OFCs. The OMIM list performed better than the Prevention Genetics or NHS lists for CP (16.2% vs. 14.8%), but these differences were not statistically different (Supplemental Table S8).

## DISCUSSION

Genetic diagnostics are currently performed on OFC cases with an OFC family history consistent with Mendelian inheritance patterns or individuals with syndromic presentations. Consequently, diagnostic testing is conducted in only a small fraction of cases, creating a potential clinical diagnostic gap. Previous OFC studies estimated a diagnostic yield of ∼10% using ES in a small set of 46 multiplex families^4^. In this sample of 841 cases from multiple populations and different family structures, we also estimated a yield of ∼10%, confirming previous studies^4,39^. Our study provided confirmation and moves beyond replication in several substantive ways. First, we showed that diagnostic yield varied significantly by OFC subtype. We observed an almost 20% diagnostic yield for CP but a nearly 7-fold lower yield in CL. The CP yield estimate was comparable to a recent report of 30 isolated CP cases, where 17% of cases had LP variants^42^. These findings could be clinically useful as they may suggest a reexamination of whether routine clinical testing of CP cases is warranted as standard of care. It is also scientifically useful as it reinforces the etiologic heterogeneity in OFC subtypes observed from studies of common variants^33,43-45^ and extends it to rare variants. It also motivates future CL-specific research which will be necessary to perform genetic testing clinically for OFCs in general. Second, we show that although some risk is clustered in nine genes, OFCs are highly heterogeneous within and between subtypes. Finally, we found that VUSs, which constitute most variants categorized in this study and in clinical tests, are enriched in cases and subsets of these VUSs, such as those in high-confidence OFC genes, are likely to contribute to the OFC phenotype.

Our estimated yield (∼10%) is similar to those from exome or genome-based studies of other pediatric conditions including congenital heart disease (12.7%)^46^ and autism spectrum disorder (7.5%)^47^. A major determining factor in these studies is their approach and cohort ascertainment. For example, Lowther and colleagues estimated the yield from sequencing in autism spectrum disorder at 7.5% using a panel of 907 neurodevelopmental genes^47^. They found a similar yield (12%) for a heterogeneous group of fetal structural anomalies in 2,535 genes but noted cases had been pre-screened by karyotype and chromosomal microarray analyses, lowering their diagnostic yield. Targeted investigations such as ours may favor specificity (but lose sensitivity) since the overall yield will be lower than exome-wide studies or including other first-tier techniques such as karyotyping or microarray. Further, diagnostic yields are typically higher in syndromic cases or those with multiple congenital anomalies^48,49^. We note that our study was not population-based so the true diagnostic yield remains to be determined.

We identified LP variants in 39 genes. Although several genes had multiple variants, few individual variants recurred, underscoring the extensive allelic and genetic heterogeneity of OFCs. We observed patterns of variation across OFC subtypes consistent with the literature. For example, we found *COL2A1* variants exclusively among CP cases^50^, *TFAP2A* variants were found exclusively among CLP cases^51^, and *IRF6* variants were found in either CP or CLP cases^52^. Interestingly, despite a strong genotype-phenotype correlation between *GRHL3* (MIM: 608317) and CP^53,54^, the only LP variant identified in *GRHL3* was in a CL case. Due to differences in sample sizes for each OFC subtype, we were unable to quantitatively analyze genotype-phenotype correlations for each gene, so these remain anecdotal observations requiring follow-up in larger datasets.

Interpreting a VUS is a considerable challenge. Nearly 62% of variants were classified as VUSs, most of which were missense variants. The effect of missense variants is often difficult to interpret without independent functional evidence, especially for genes with high allelic heterogeneity. There were multiple genes (e.g., *PRICKLE1*) for which VUSs were identified in cases while zero VUSs were identified in controls. *PRICKLE1* was previously evaluated through family-based association studies and showed evidence of association with OFCs^55^. Here, we found a similar, but nominally significant, effect on OFC risk. Therefore, it is likely these datasets are underpowered to detect genes with a burden of rare variants and the top-ranked genes should be considered for further analysis pending functional testing to sort out the effect of identified variants.

It is important to note this study was conducted on a cross-sectional cohort from multiple recruitment protocols intended for research, and it is not representative of all OFC cases that may be referred from craniofacial clinics. Clinical diagnostics and differential diagnoses are aided by detailed phenotyping and collection of family histories, but data availability is limited for specific populations in this cohort. Although those with multiple congenital anomalies and significant developmental delays were likely excluded and such individuals should represent a minority of the dataset, the recruitment timing, varying skills of the clinical and research teams, and different recruitment goals make this a highly heterogeneous cohort with incomplete phenotypic data needed for the clinical setting. However, many OFC syndromes show incomplete penetrance and variable expressivity, which can complicate a diagnosis based on phenotype alone even when detailed phenotyping is available. In this study, we estimated the penetrance of OFCs for transmitted alleles but could not estimate the extent of variable expressivity of other phenotypic features. Moreover, these estimates represent global penetrance, not gene-level, which requires additional investigation in larger cohorts. Nonetheless, the low penetrance of LP variants was striking, as many variants were predicted to be ‘loss of function’. The ideal cohort to fully evaluate penetrance and expressivity would be a prospectively recruited, deeply phenotyped, and sequenced cohort of sequential cases, which are difficult and costly to assemble. Lastly, we only reviewed some variants to limit variant “noise”; however, some variants that did not meet our prioritization criteria could be disease-causing.

Sequencing studies such as this one and those investigating the functional consequences of variants in model systems are necessary to advance research translation to clinical practice. Although panel-based clinical tests will inevitably be replaced by GS, our results offer insight into the breadth of genes that may be found from clinical sequencing. Our results suggest there is value in genetic testing of CP cases but limited utility in using panels for CL cases. It remains to be seen if the yield differences are due to differences in architecture (i.e. fewer highly penetrant variants in CL) or differences in panel content that are insufficient for CL. Besides the potential clinical applications, we highlight the critical need for high-throughput validation to quantitatively distinguish the effects of individual rare variants. Future work in this area should allow for improved variant interpretation in a clinical setting, a greater understanding of the genes influencing craniofacial birth defects, and help explain the variable penetrance observed in this study.

## Data Availability

The case data analyzed and reported in this manuscript were accessed from the database of Genotypes and Phenotypes (dbGaP; European trios, dbGaP: phs001168.v2.p2; Colombian trios, dbGaP: phs001420.v1.p1; Taiwanese trios, dbGaP: phs000094.v1.p1) and from the Kids First Data Resource Center. The control data is available from public data repositories as described in https://www.internationalgenome.org/data-portal/data-collection/30x-grch38.

## Data Availability

The case data, including cleft subtype and associated genome sequencing data, analyzed and reported in this manuscript were accessed from the database of Genotypes and Phenotypes (dbGaP; European trios, dbGaP: phs001168.v2.p2; Colombian trios, dbGaP: phs001420.v1.p1; Taiwanese trios, dbGaP: phs000094.v1.p1) and from the Kids First Data Resource Center. The control data is available from public data repositories as described in https://www.internationalgenome.org/data-portal/data-collection/30x-grch38. Sample IDs in the supplemental data are the same as the “Case IDs” on the Kids First Cavatica data portal.

## Acknowledgements

This study would not be possible without the dedication of many families, study teams, and colleagues worldwide.

## Author Contributions

Conceptualization: EJL; Data curation: KKDP, EJL, SWC, MRB, AS-J, XZ, HB, TM; Formal analysis: KKDP, EJL, SH, BC, AS-J, TH, ; Funding acquisition: EJL, MLM, SMW, EF, THB, HB, KKDP; Methodology: KKDP, DJC, MPE, HB, EJL; Resources: LCV-R, CR, JTH, LMU, GW, SMW, THB, JCM, EF, MLM, HB; Software: AS-J, XZ, HB; Writing-initial draft: KKDP, EJL; All authors critically reviewed, edited, and/or approved the final manuscript.

