## Supplemental Figure for "Rare variants found in clinical gene panels illuminate the genetic and allelic architecture of orofacial clefting"

**SUPPLEMENTAL FIGURES**

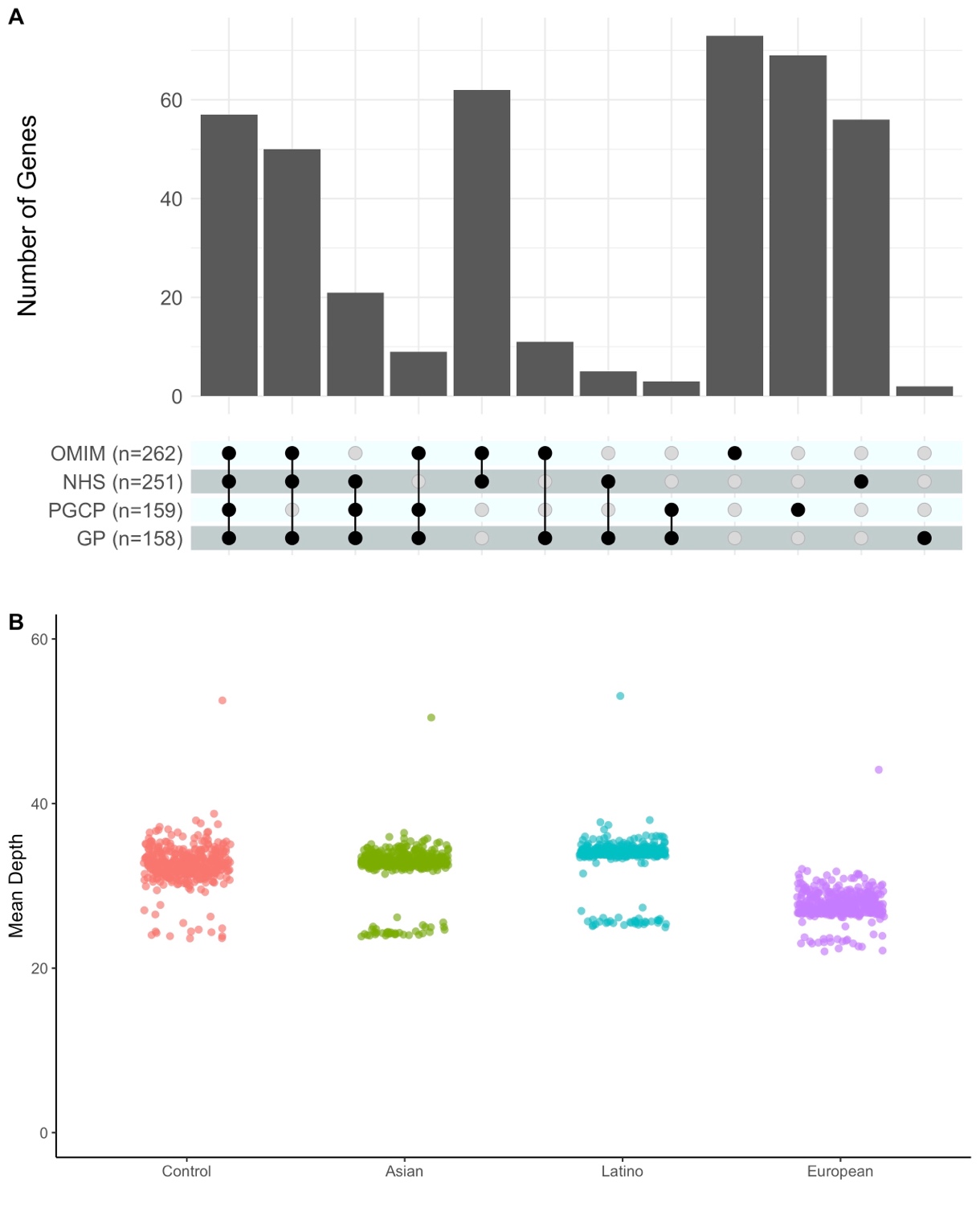

**Figure S1. OFC Gene List.** **(A)** 418 genes associated with OFCs and craniofacial development obtained from four sources: the Online Mendelian Inheritance in Man (OMIM), the National Health Service (NHS), Prevention Genetics Cleft Lip & Palate Panel (PGCP), and a manually curated gene panel (GP). The overlap of genes between sources is shown by the filled circles and connecting lines on the lower panel while the number of genes in each overlap and individual source is shown by the gray bars. **(B)** Sequencing coverage of the OFC genes (circles) by study population: Controls (pink), Asian (green), Latino (blue), and European (purple).

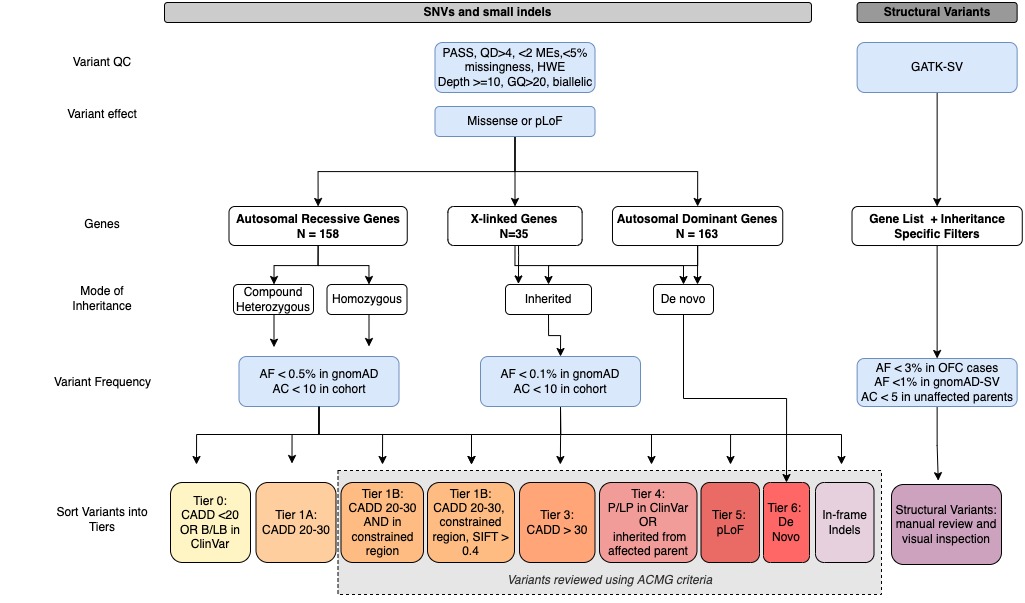

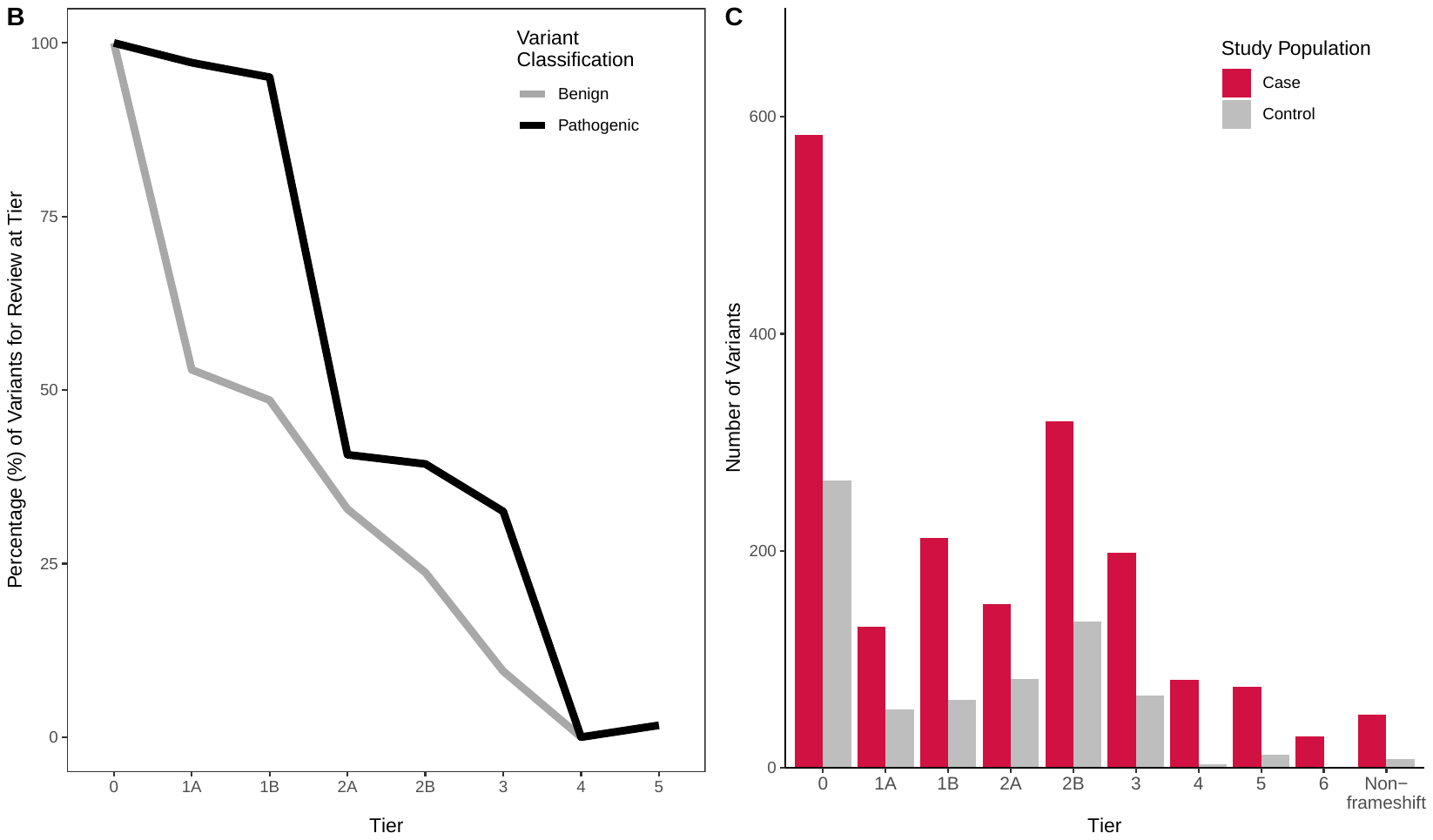

**A**

**Figure S2.** **Whole-Genome Sequencing and Variant Filtering Pipeline.** (A) Pipeline for prioritizing and sorting single-nucleotide variants (SNVS), in-frame insertions and deletions, and structural variants. (B) Percentage of pathogenic (black) and benign (gray) ClinVar variants in genes from the OFC gene list at each tier threshold for Tiers 0-5. (C) The number of variants from 841 OFC cases (red) and 294 controls (grey) sorted into tiers.

CADD: Combined Annotation Dependent Depletion; B/LB: ‘Benign’ or ‘Likely Benign’ Variants; SIFT: Sorting Intolerant from Tolerant; P/LP: ‘Pathogenic’ or ‘Likely Pathogenic’ Variants; pLoF: Predicted Loss-of-Function Variants

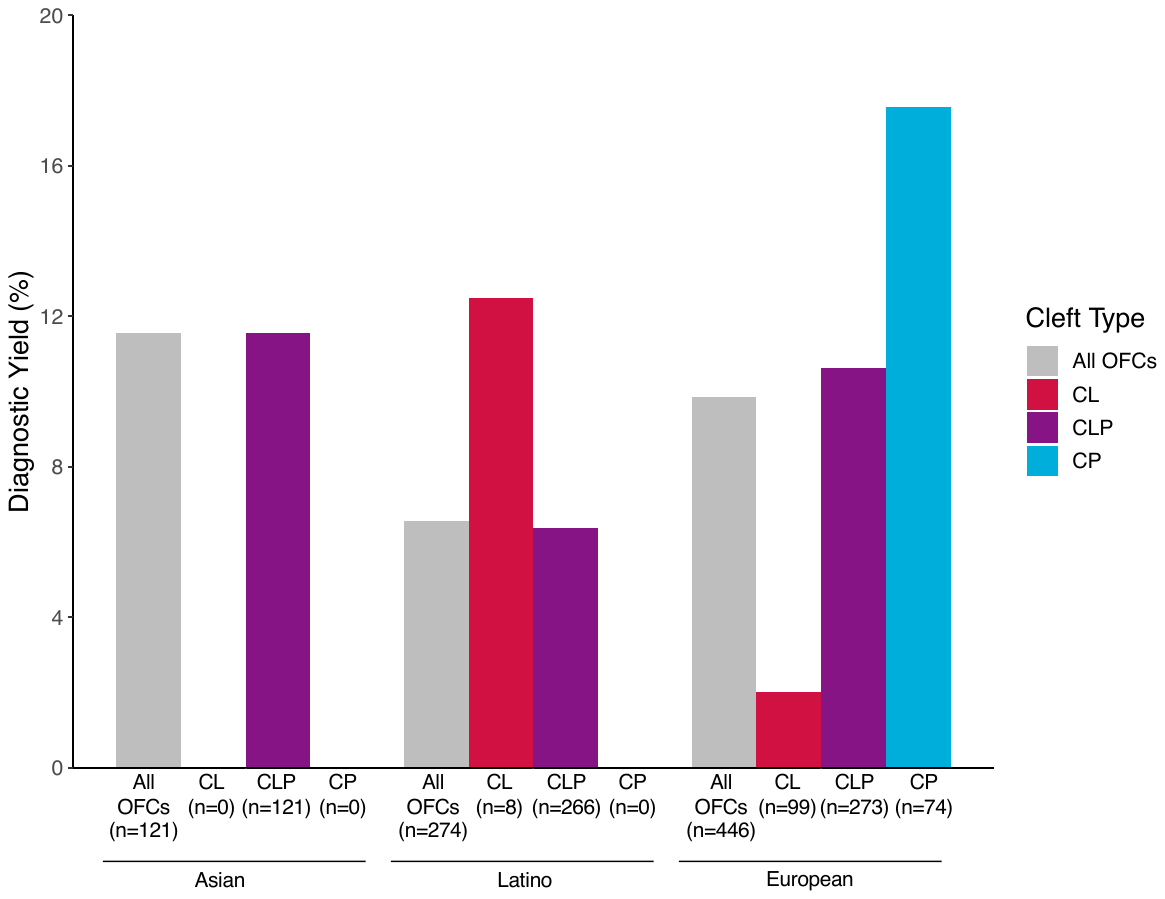

**Figure S3. Diagnostic Yield by Cleft Type and Population.** The percentage of cases with ‘likely pathogenic’ variants are shown by cleft type, including all OFCs (gray), CL (red), CLP (purple), and CP (blue) and population. The sample size for each OFC subtype and population group is denoted below each bar.

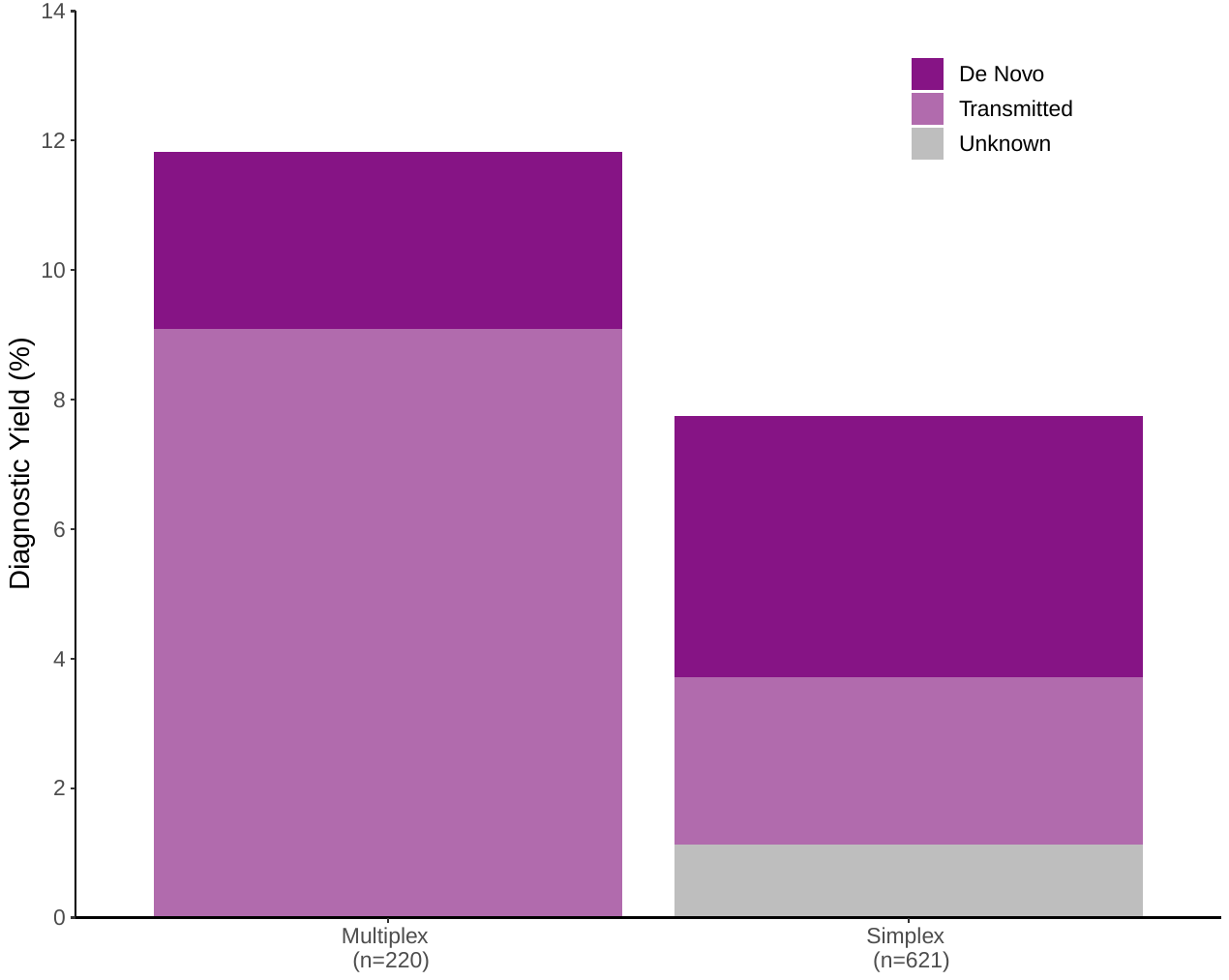

**Figure S4. Diagnostic Yield by Family Type.** The diagnostic yield from multiplex families (N=220) versus simplex families (N=621). ‘Likely pathogenic’ variants are classified by mode of inheritance: *de novo* (dark purple), transmitted from a parent (light purple), or unknown (gray).

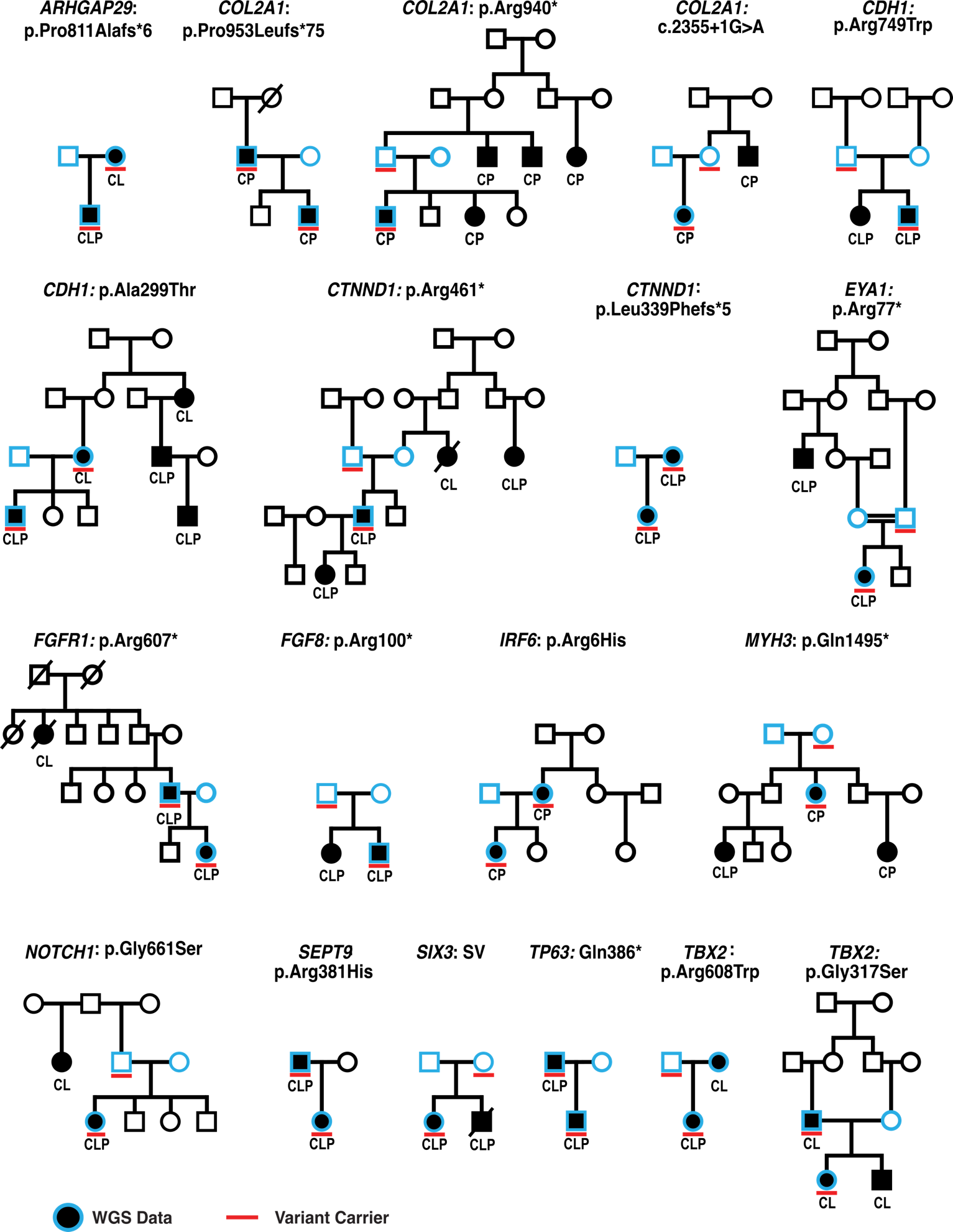

**Figure S5. Inherited Likely Pathogenic Variants in Multiplex Families.** The mutated gene and consequence are noted above each pedigree (refer to Table S6 for variant details). Sex symbols with solid black indicate the phenotype of the individual: CL (cleft lip), CP (cleft palate), and CLP (cleft lip and palate). The red solid lines below individuals indicate variant carriers while the blue outline of the sex symbols indicates individuals with WGS data.

Genes and protein identifiers: *ARHGAP29* (NP_004806.3); *COL2A1* (NP_001835.3); *CDH1* (NP_004351.1); *CTNND1* (NP_001322.1); *EYA1* (NP_000494.2); *FGFR1* (NP_001167534.1); *FGF8* (NP_006110.1); *IRF6* (NP_006138.1); *MYH3* (NP_002461.2); *NOTCH1* (NP_060087.3); *SEPT9* (NP_006631.2); *TP63* (NP_003713.3); *TBX2* (NP_005985.3)

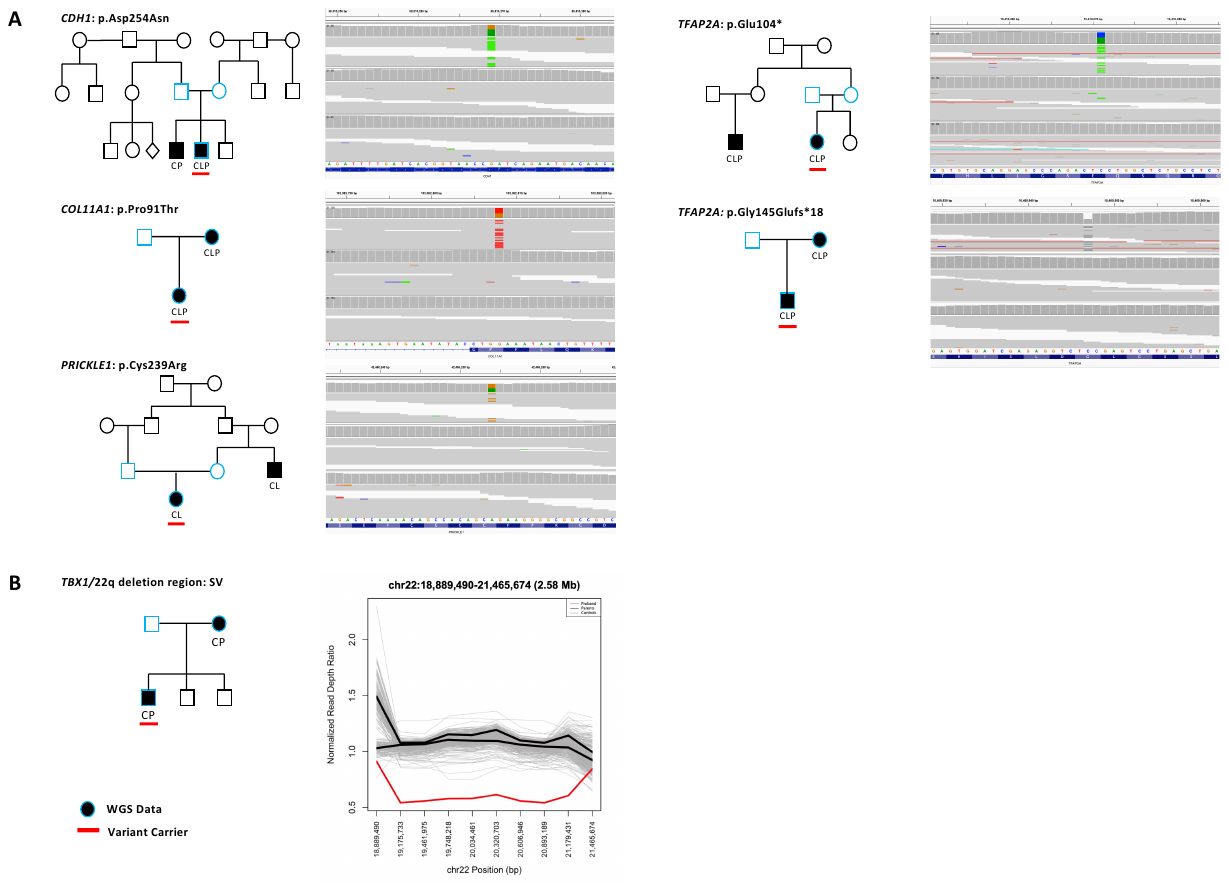

**Figure S6. *De Novo* Mutations in Multiplex Families. (A)** We identified ‘likely pathogenic’ *de novo* variants in *CDH1*, *COL11A1*, *PRICKLE1*, and *TFAP2A* (refer to Table S6 for variant details). We confirmed the *de novo* mutation by visual inspection of the proband (top segment), father (middle segment), and mother (bottom segment) reads using the Integrative Genomics Viewer (IGV) ^38^. **(B)** We identified a *de novo* deletion encompassing the *TBX1*/22q deletion region. On the right, we the read depth ratio of the proband (red), parents (black), and controls (gray) of the SV region on the right. In each pedigree, the red solid lines below symbols indicate variant carriers while the blue outline correspond to sequenced individuals.

Genes and protein identifiers: *CDH1* (NP_004351.1); *COL11A1* (NP_542197.3); *PRICKLE1* (NP_001138353.1); *TFAP2A* (NP_003211.1)

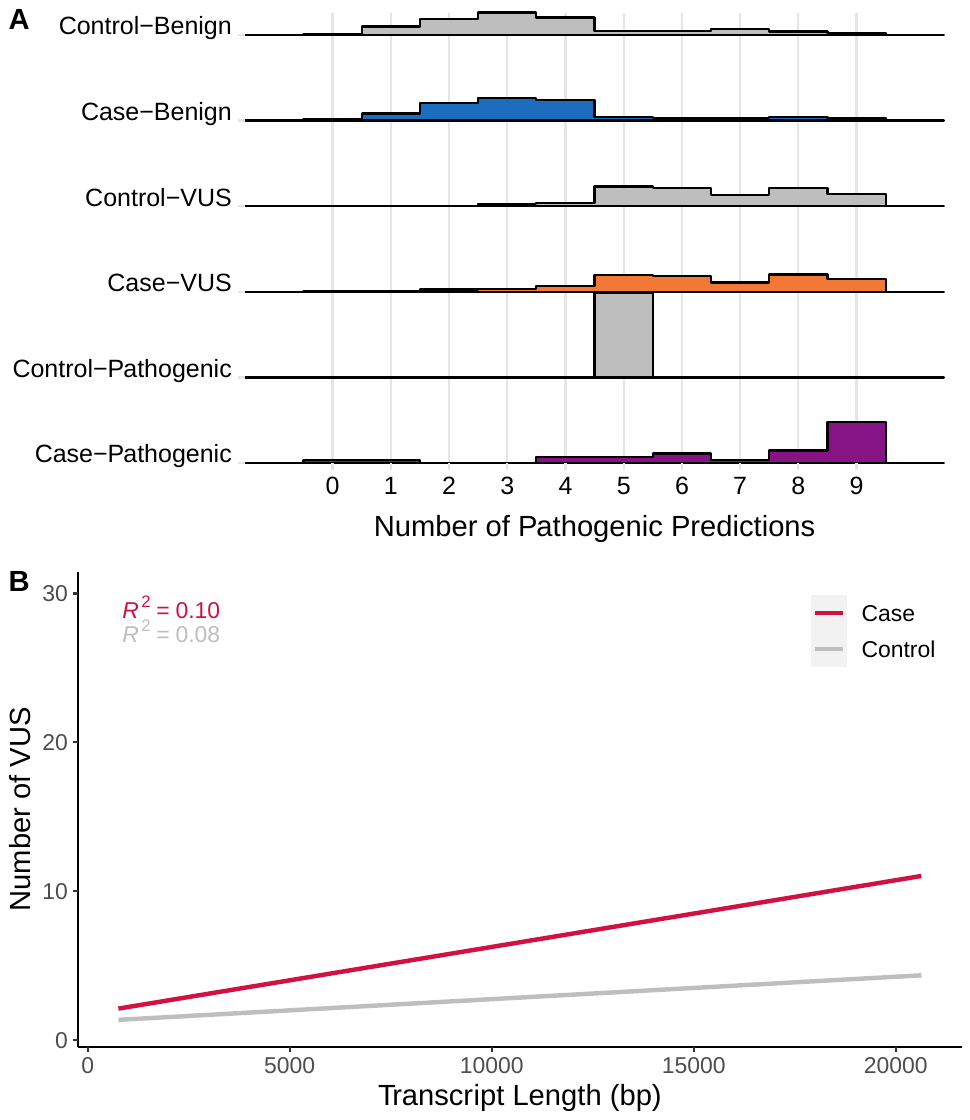

**Figure S7**. **Variants of Uncertain Significance in Cases and Controls.** (A) Distribution of the number of *in silico* prediction tools from nine different algorithms predicting a missense variant to be likely pathogenic/damaging for ‘likely benign’, VUS, and ‘likely pathogenic’ variants. (B) The number of VUS in genes is correlated with transcript length in cases (red, p=1.28 x 10^-05^) and controls (gray, p=3.82 x 10^-03^).

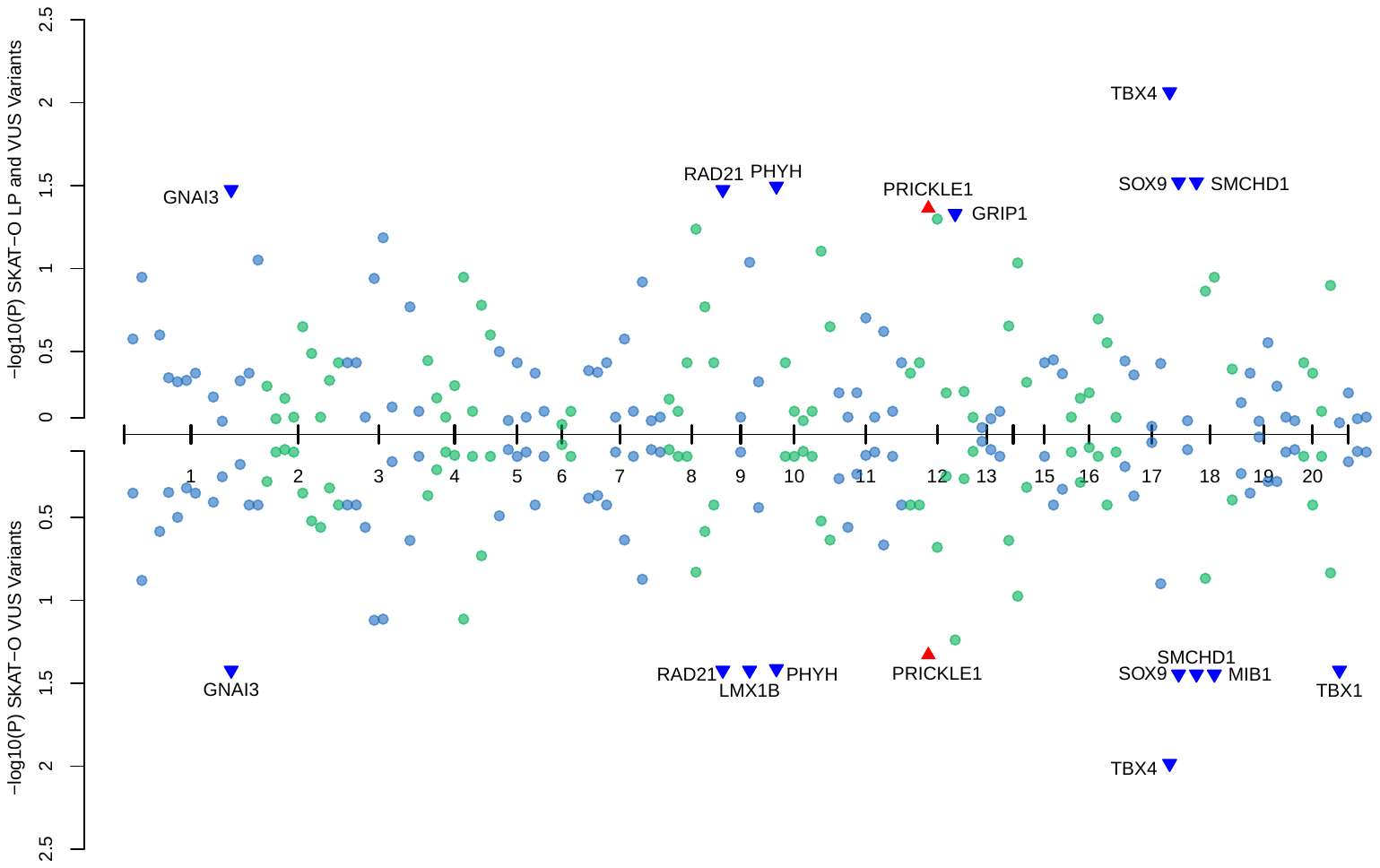

**Figure S8. Gene-Based Association Tests of VUS**. SKAT-O gene-based association tests for 139 genes with VUS and/or ‘likely pathogenic’ (LP) variants (top) and 129 genes with VUS variants (excluding individuals with ‘likely pathogenic’ variants) (bottom). Each gene with p < 0.05 is labeled according to the direction of effect with a triangle: decreased risk for cases (dark blue) and increased risk for cases (red). No gene reached a formal Bonferroni significance threshold (p < 3.60 x 10^-4^ (top) and p < 3.88 x 10^-4^ (bottom)).

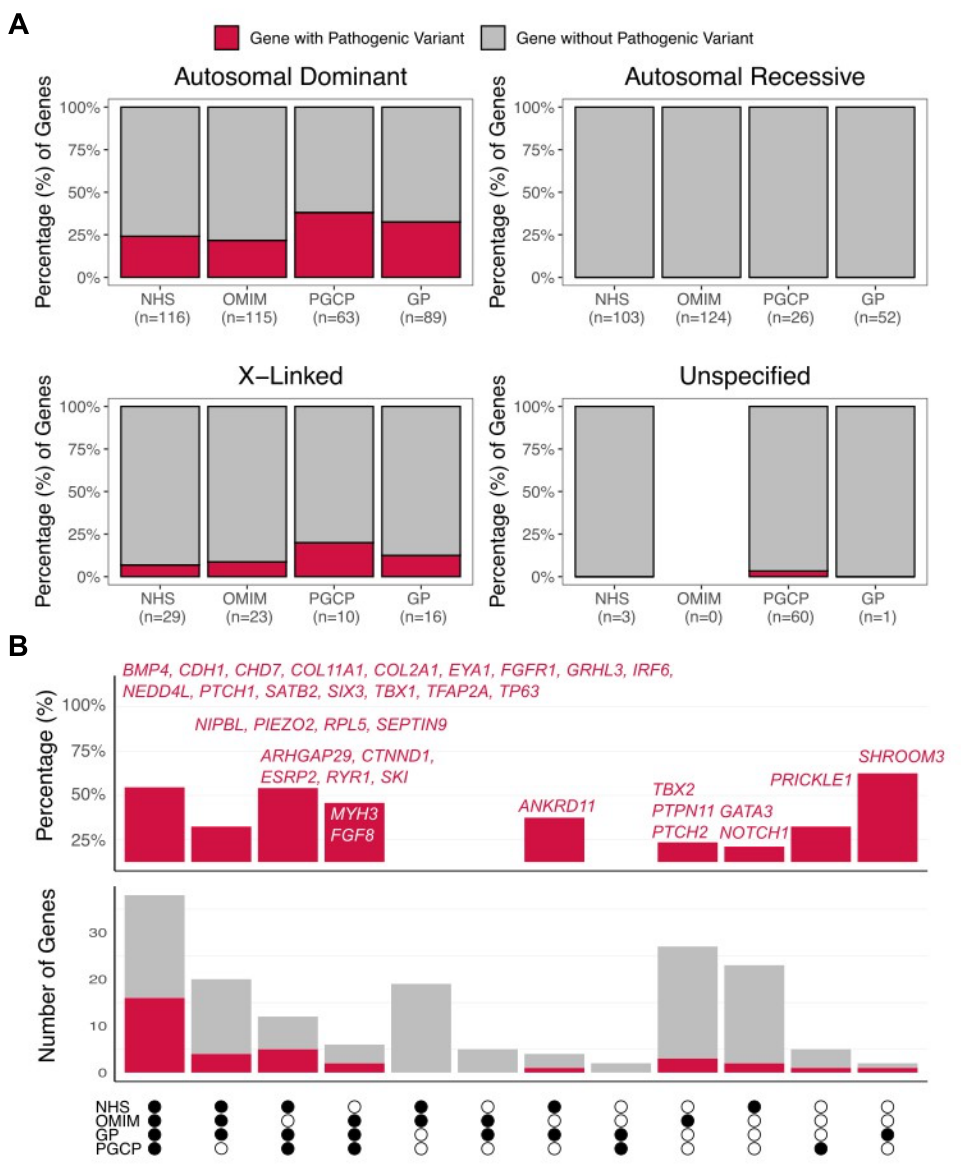

**Figure S9. Genes with Likely Pathogenic Variants by Gene List Source.** (A) The proportion of autosomal dominant, autosomal recessive, X-linked, and unspecified genes from each source list with ‘likely pathogenic’ variants (red). (B) The top axis shows the percentage of autosomal dominant genes in each source list overlap set with ‘likely pathogenic’ variants; genes with these variants are listed. The bottom axis shows the number of genes (red) with ‘likely pathogenic’ variants and the number of genes in the overlap set without ‘likely pathognic’ variants (grey). The overlap sets are indicated on the x-axis by filled circles for each source list that a gene was on: GP (curated gene panel), NHS (NHS PanelApp list), OMIM, and PGCP (Prevention Genetics panel).

**SUPPLEMENTAL TABLES**

**Table S1. Demographics of OFC Cohort**

**Table S2. Family History Information for Study Populations**

**Table S3. OFC Gene List**

**Table S4. Analyzed Structural Variants**

**Table S5. American College of Medical Genetics & Genomics Classification Modifications**

**Table S6. Likely Pathogenic Variants in Present Study**

**Table S7. VUS Comparisons by Population and Cleft Type**

**Table S8. Yield by Cleft Type and Source Gene List**
